# Investigation of the sero-epidemiology of vaccine preventable diseases and common viral infections in French populations using a multiplex serological assay

**DOI:** 10.1101/2024.04.26.24306413

**Authors:** Emma Bloch, Gaëlle Baudemont, Françoise Donnadieu, Laura Garcia, Stéphane Pelleau, SeroPed Study Consortium, Milieu Intérieur Consortium, Lluis Quintana-Murci, Darragh Duffy, Arnaud Fontanet, Michael White

## Abstract

Vaccine-preventable diseases (VPDs) and common viral infections (CVIs) pose substantial public health challenges. Despite data collected through routine vaccination programs and through hospital-based case reporting systems, the understanding of population-level immunity to VPDs and CVIs remains limited. To address this challenge, this study focuses on the development and application of a high-throughput multiplex serological assay to assess IgG levels against a large panel of eight VPDs (e.g., tetanus, diphtheria, measles, mumps, rubella) and 18 CVIs that typically occur in early childhood (e.g., adenovirus, cytomegalovirus, respiratory syncytial virus). A cross-sectional study of 2,032 serum samples from French children and adults was used to optimize and validate a 47-plex assay. These antibody measurements allowed us to enhance our understanding of immunity, vaccination and booster dose effect, as well as antibody waning over time. Preliminary results demonstrate the importance of age-stratified IgG level measurements, revealing notable variation in seroprevalence patterns among different age groups. These findings suggest that there are variations in immunity and exposure to the targeted pathogens across these groups. These findings highlight the potential of high-throughput multiplex serology assays for routine surveillance of common viral infections and assessment of vaccine coverage.

## Introduction

Serology is used for surveillance by detecting past or concurrent infections by measuring antibodies to specific pathogens. Traditionally, serological tests have been based on assaying the functional activity of antibodies, for example by observing how antibodies bind and neutralize a virus and prevent the formation of viral plaques. At present, the most widely used serological test is an enzyme-linked immunosorbent assay (ELISA) which measures antibodies bound to a target antigen. Neutralization assays and ELISA assays are single-plex, meaning that each experiment measures one biomarker.

Multiplex platforms allow antibody responses to multiple antigens to be measured using as little as 1 μL of serum. Notable technologies include bead-based assays (10s – 100s of antigens), protein microarrays (100s – 1,000s of antigens)^1^ and phage arrays (100,000+ antigens)^2^. Two of the most widely used multiplex platforms for serological assays are the Luminex MAGPIX^®^ (capable of measuring 50 analytes per sample) and the Luminex INTELLIFLEX^®^ (500 analytes per sample). Advantages of these bead-based systems are low cost per sample, low sample volume, low measurement error, and ease of transfer to other labs.^3–5^ Challenges to the development of multiplex serological assays include difficulties in identifying experimental conditions that are optimal for all considered pathogens, selection of appropriate controls, and analysis of the large amount of data generated.

Two of the most promising applications for multiplex assays are serological surveillance of vaccine preventable diseases (VPDs) and common viral infections (CVI). Vaccines targeting a variety of pathogens, including viruses and bacteria, have been introduced to prevent the occurrence of diseases like tetanus, diphtheria, measles, mumps, rubella, whooping cough, and human papillomavirus (HPV)^6^. These programs have been essential in reducing the incidence of VPDs and alleviating their societal burden. The coverage of vaccination programs can be estimated from national public health data, but this data is typically incomplete. Vaccination coverage can instead be accurately estimated from serological surveys based on random samples from the general population. A notable example is the case of The Netherlands where multiplex serological assays are routinely used for surveillance of vaccine coverage^7^.

CVIs such as Adenovirus, Respiratory Syncytial Virus (RSV), Norovius, seasonal Coronavirus and Enteroviruses, pose a persistent burden to human health and can cause various respiratory and gastrointestinal symptoms. While these infections are generally mild, they can lead to more severe complications in children and immunocompromised individuals^8–10^. Additionally, the ongoing emergence of new infectious agents such as SARS-CoV-2 underscores the need for continuous surveillance to address emerging infectious threats^11–13^.

To gain insights into circulating pathogens and assess population protection against infections, a focus on pathogen surveillance and immune monitoring is crucial. Valuable information about prevalent respiratory viruses and seasonal patterns are provided by routine PCR testing of hospitalized patients with clinical symptoms^12,14^. However, its effectiveness as a surveillance tool is constrained by the large number of asymptomatic or mild symptomatic respiratory infections^15^. This limitation makes it impractical to estimate the prevalence of infections solely through PCR testing, particularly for common viral infections. Additionally, the abundance of co-circulating pathogens, sharing similar clinical profiles, notably fever and cough, poses a significant challenge for symptomatic surveillance, particularly in environments where children gather, such as daycare and school settings^16–18^. This study specifically focuses on immune surveillance, employing antibody measurements as a cost-effective method to assess seroprevalence and immunity in the population.

Understanding both individual and population-level immunity is essential for designing effective surveillance programs. Individual immunity, acquired through natural infection or vaccination, is influenced by various factors such as age, genetics, and environmental exposures. In contrast, population immunity, also known as herd immunity, refers to the indirect protection against infectious diseases that occurs when a substantial portion of the population become immune, either through vaccination or prior infection. This concept plays a crucial role in limiting the spread of infectious agents within a community.

In response to an infection or vaccination, the immune system produces antibodies, also known as immunoglobulins (Ig), that possess the ability to recognize and neutralize a pathogen. Among these, IgG antibodies persist for years after infection or immunization and are actively synthesized by plasma B cells, which originate from the differentiation of memory B cells^19^. This ensures a continuous and sustained production of antibodies, contributing significantly to long-term immunity.^7^ This robust immune response prevents reinfections and mitigates the severity of the disease in the event of a second infection. In the case of children, monitoring antibody levels provides valuable insights into their first exposure to an infectious disease and vaccination status.

Here we describe the development of a high-throughput multiplex serology assay to assess IgG levels against a comprehensive panel of eight vaccine-preventable diseases (VPDs) comprising tetanus, diphtheria, measles, mumps, rubella, whooping cough, papillomavirus, and 18 common childhood viral infections (CVIs) typically occurring in early childhood stages, including Adenovirus, Cytomegalovirus (CMV), RSV and Varicella-Zoster Virus (VZV). This assay is applied to samples from the French population to provide a quantitative description of multi-pathogen sero-epidemiology.

## Material and Methods

### Study population and samples

#### The Seroped cohort

In this cross-sectional study 1,132 serum samples were collected from children and adults who were attending hospitals in northeastern France between February 2020 and August 2020 to evaluate the seroprevalence of SARS-CoV-2^20^. The samples used in this study consisted of leftover samples derived from routine medical testing within French hospital laboratories. They were processed in accordance with existing regulations and guidelines of the French Commission for Data Protection (Commission Nationale de l’Informatique et des Libertes). All identifiable information was destroyed for these samples, with the exception of age and sex.

Table 1 summarizes the number and percentage of individuals in each age and sex group. The age of the sampled individuals ranges from 0 to 100 years old, with a special note for those labeled <1 who were sampled at the hospital at the time of birth. This last inclusion allows us to gain insights into the presence of maternal antibodies

**Table 1:** Demographic characteristics of 1,132 subject sampled in French Hospitals in 2020.

|  | count (N) | percent (%) |
| --- | --- | --- |
| Age(years): |  |  |
| <1 | 19 | 1.7 |
| 1 | 29 | 2.6 |
| 2 | 20 | 1.8 |
| 3 | 24 | 2.1 |
| 4 | 31 | 2.7 |
| 5 | 29 | 2.6 |
| 6 | 26 | 2.3 |
| 7 | 25 | 2.2 |
| 8 | 42 | 3.7 |
| 9 | 18 | 1.6 |
| 10 | 31 | 2.7 |
| 11-12 | 61 | 5.4 |
| 13-14 | 64 | 5.7 |
| 15-17 | 179 | 15.8 |
| 18-29 | 167 | 14.8 |
| 30-45 | 75 | 6.6 |
| 46-60 | 98 | 8.7 |
| >60 | 194 | 17.1 |
| Sex: |  |  |
| Male | 550 | 48.6 |
| Female | 488 | 43.1 |
| Unknown | 94 | 8.3 |
| Total: | 1,132 |  |

#### The Milieu Intérieur cohort

The *Milieu Intérieur* (MI) cohort consists of 1,000 healthy adults recruited in Rennes, France between 2012 and 2013^21^. Donors were chosen based on the absence of any indication of severe, chronic, or recurrent medical conditions. Samples from 900 individuals were available for the current study, 453 female and 447 male, ranging from 20 to 70 years of age. This cohort is used to validate our assay by comparing it with serological data generated using clinically graded immunoassays.

The clinical study was approved by the Comité de Protection des Personnes - Ouest 6 on June 13, 2012, and by the French Agence Nationale de Sécurité du Médicament on June 22nd, 2012. The study is sponsored by Institut Pasteur (Pasteur ID-RCB Number: 2012-A00238-35) and was conducted as a single-center study without any investigational product. The original protocol was registered under ClinicalTrials.gov (study# NCT01699893). The samples and data used in this study were formally established as the *Milieu Intérieur* biocollection (NCT03905993), with approvals by the *Comité de Protection des Personnes – Sud Méditerranée* and the *Commission Nationale de l’Informatique et des Libertés* (CNIL) on April 11, 2018.

### Antigens-Recombinant proteins

We assembled a list of antigens for pathogens of epidemiological interest, encompassing a total of 15 distinct antigens targeting eight vaccine-preventable diseases (Tetanus, Diphtheria, Measles, Mumps, Rubella, Human papillomavirus, Whooping cough) and 32 antigens directed against 15 viral infections commonly observed during early childhood stages (including Adenovirus, Cytomegalovirus, Respiratory syncytial virus).

The proteins used were either purchased from Native Antigen (Oxford, UK), ProSpec-Tany Techno Gene (Israel), Ray Biotech (Georgia, US) or NIBSC (Herts, UK). Antigen names, expression systems, suppliers, catalogue numbers, along with coupling conditions (optimal antigen concertation and coupling buffer) are provided in Supplementary Table 1.

**Table 2:** List of antigen names and suppliers used for assay development.

| Antigen name | Short name | Supplier | Catalog number |
| --- | --- | --- | --- |
| *Bordetella pertussis*. Toxin | Bordetella p. Toxin | The Native Antigen | PT-TNL-50 |
| *Bordetella pertussis*, Filamentous haemagglutinin | Bordetella p. FHA | The Native Antigen | BP-FHA-50 |
| *Corynebacterium diphtheriae* toxin | Diphtheria Toxin | The Native Antigen | DIP-TNL-100 |
| Tetanus Toxin | Tetanus Toxin (NA) | The Native Antigen | REC31801-100 |
| Tetanus Toxin | Tetanus toxin | NIBSC | 02/232 |
| Measles virus lysate | Measles lysate | The Native Antigen | NAT41576-100 |
| Measles virus nucleoprotein | Measles nucleoprotein | The Native Antigen | REC31796-100 |
| Mumps lysate | Mumps lysate | The Native Antigen | NAT41577-100 |
| Mumps virus nucleoprotein | Mumps nucleoprotein (PS) | ProSpec-Tany TechnoGene | MMP-001 |
| Mumps virus nucleoprotein | Mumps nucleoprotein | The Native Antigen | REC31810-100 |
| Rubella virus-like particles | Rubella VLP | The Native Antigen | REC31651-100 |
| Rubella virus Capsid | Rubella Capsid | ProSpec-Tany TechnoGene | RUB-293 |
| Rubella virus E2 | Rubella E2 | ProSpec-Tany TechnoGene | RUB-292 |
| Hepatitis B Virus E antigen | Hepatitis B E antigen | The Native Antigen | REC31677-100 |
| Hepatitis B Virus Core antigen | Hepatitis B core antigen | The Native Antigen | REC31689-100 |
| Hepatitis B Virus Surface antigen | Hepatitis B surface antigen | ProSpec-Tany TechnoGene | HBS-872 |
| Human Papillomavirus serotype 16 | HPV 16 | ProSpec-Tany TechnoGene | HPV-001 |
| Human Papillomavirus serotype 18 | HPV 18 | ProSpec-Tany TechnoGene | HPV-002 |
| Varicella-Zoster Virus, Heterodimer gE/Gi | Varicella-Zoster Virus | The Native Antigen | REC31907-100 |
| Adenovirus T3 | Adenovirus T3 lysate | The Native Antigen | AD004-100 |
| Adenovirus T5 | Adenovirus T5 lysate | The Native Antigen | AD005-100 |
| Adenovirus Type 5 Hexon Protein | Adenovirus T5 | The Native Antigen | AH01-100 |
| Adenovirus Type 40 Hexon Protein | Adenovirus T40 | The Native Antigen | NAT41552-100 |
| Cytomegalovirus | Cytomegalovirus | The Native Antigen | CMV-HP-100 |
| Espein Barr Virus gp125 | Espien-Barr virus | The Native Antigen | REC31601-100 |
| Echovirus | Echovirus | The Native Antigen | REC31776-100 |
| Enterovirus CoxB3 VP1 | Enterovirus CoxB3 VP1 | The Native Antigen | REC31738-10 |
| Hepatitis A Virus | Hepatitis A | RayBiotech | 227-10025 |
| Hepatitis C Core antigen | Hepatitis C Core antigen | The Native Antigen | REC31693-100 |
| Hepatitis E Virus ORF2 | Hepatitis E ORF2 | The Native Antigen | REC31653-100 |
| Norovirus GII.4 VP1 | Norovirus GII.4 VP1 | The Native Antigen | REC31620-100 |
| Norovirus GII.6 VLP | Norovirus GII.6 | The Native Antigen | REC31985-100 |
| Respiratory Syncytial A | Respiratory Syncytial A | The Native Antigen | NAT41624-100 |
| Respiratory Syncytial B | Respiratory Syncytial B | The Native Antigen | NAT41625-100 |
| RSV gG | Respiratory Syncytial gG | The Native Antigen | RSV-GPB-50 |
| Rhinovirus T1A lysate | Rhinovirus T1A lysate | The Native Antigen | NAT41626-100 |
| Rotavirus VP7 | Rotavirus VP7 | The Native Antigen | REC31910-100 |
| Toxoplasma gondii SAG2 | Toxoplasma gondii SAG2 | The Native Antigen | REC31825-100 |
| Human coronavirus OC43 nucleoprotein | OC43 nucleoprotein | The Native Antigen | REC31857-100 |
| Human coronavirus OC43 Spike protein | OC43 spike | The Native Antigen | REC31894-100 |
| Human coronavirus HKU1 nucleoprotein | HKU1 nucleoprotein | The Native Antigen | REC31856-100 |
| Human coronavirus HKU1 Spike protein | HKU1 spike | The Native Antigen | REC31897-100 |
| Human coronavirus 229E nucleoprotein | 229E nucleoprotein | The Native Antigen | REC31758-100 |
| Human coronavirus 229E Spike protein | 229E spike | The Native Antigen | REC31895-100 |
| Human coronavirus NL63 nucleoprotein | NL63 nucleoprotein | The Native Antigen | REC31759-100 |
| Human coronavirus NL63 Spike protein | NL63 spike | The Native Antigen | REC31896-100 |
| Influenza A H1N1 Hemagglutinin | Influenzavirus A | The Native Antigen | FLUH1N1-HA-100 |

We assembled a list of antigens for pathogens of epidemiological interest, encompassing a total of 15 distinct antigens targeting eight vaccine-preventable diseases (Tetanus, Diphtheria, Measles, Mumps, Rubella, Human papillomavirus, Whooping cough) and 32 antigens directed against 15 viral infections commonly observed during early childhood stages (including Adenovirus, Cytomegalovirus, Respiratory syncytial virus).

The proteins used were either purchased from Native Antigen (Oxford, UK), ProSpec-Tany Techno Gene (Israel), Ray Biotech (Georgia, US) or NIBSC (Herts, UK). Antigen names, expression systems, suppliers, catalogue numbers, along with coupling conditions (optimal antigen concertation and coupling buffer) are provided in Supplementary Table 1.

### Antigen-bead coupling

To establish this assay, 47 distinct Luminex magnetic bead regions were utilized, where each antigen was coupled to a specific bead type. The magnetic beads were vortexed before being transferred to a 1.5mL microcentrifuge tube. For the preparation of a 500 µL antigen-coupled bead premix, 125 µL of each Luminex magnetic bead type was employed. Subsequently, a magnetic rack was used to remove the supernatant before washing the beads with Milli-Q water. The beads were activated using 0.1M sodium phosphate (NaP) pH 6.2, 10 mg/mL of EDC (1-ethyl-3-[3-dimethylaminutesopropyl] carbodiimide hydrochloride) and 10 mg/mL sulfo-NHS (sulfo N-hydroxylsulfosuccinimide), followed by incubation on a rotor in the dark for 20 minutes at room temperature. This activation step allows the coupling of antigens to magnetic beads, through the formation of a covalent bond formed between a stable ester on the surface of the bead and the primary amine of the antigen. Thereafter, a magnetic rack was used to remove the supernatant before the beads underwent two washes with PBS 1X (phosphate-buffered saline). The mass of proteins coupled onto the beads was optimized previously, using a reference pool and validated by generating a log-linear standard curve, ensuring optimal antigen concentration coupling. The antigen-coupled beads were incubated either with PBS 1X or MES buffer (determined during the optimization stage) on a rotor in the dark overnight, at 4 °C. Subsequently, the antigen-coupled beads underwent three washes with PBS-TBN (phosphate buffer saline supplemented with 1% bovine serum albumin, 0.02% sodium acid and 0.05% Tween-20) and were resuspended in 500 µl of PBS-TBN buffer before being stored at 4 °C.

### Multiplex Serological assay

The multiplex serological assay was performed using a panel of 47 antigen-coupled beads encompassing vaccine-preventable disease and common viral infections. To measure the antibody levels against these antigens, we employed the Intelliflex® technology from Luminex®.

In a 96-well plate, 50 µL of 1/100 diluted serum (for 1/200 final serum dilution) and 50 µL of antigen-coupled beads premix (500 beads/region/µL) were added. All serum dilutions and bead premixes were prepared in PBT buffer (phosphate buffered saline containing 1% bovine serum albumin and 0,05% (v/v) Tween-20). Then the plate was incubated on a shaker in the dark for 30 min, at room temperature. Three wash steps follow incubation, placing the 96-well plate on a magnetic plate separator (Luminex®, Austin, Texas, USA) with washes performed with PBT buffer. For the detection of specific IgG, a secondary antibody conjugated to R-phycoerythrin (from Jackson Immunoresearch, UK; cat#709-116-098) was utilized at a 1/120 dilution. 50 µL of the secondary antibody was added into each well and incubated on a shaker in the dark for 15 min at room temperature. Following incubation, three wash steps were performed using a magnetic plate separator and PBT buffer. Finally, 100 µL of PBT buffer was added as the final volume. Plates were read using the Intelliflex® system at low PMT setting and the median fluorescence intensity (MFI) was measured.

Each 96-well plate included a blank (beads without serum) as a control for background signal, along with a 7 point-2-fold serum dilution (1/50 to 1/102,400) of a reference pool. During the optimization phase, a sample set of French serum samples were screened from which 15 serum samples with the highest MFI for a large number of the pathogens included in the panel were selected for the reference pool. The preliminary analysis relied on the median MFI extracted from the raw data. To conduct further analysis and account for interassay variation, a five-parameter logistic curve was employed to convert the MFI to RAU, relative to the standard curve conducted on the same plate. Microsoft Excel 2016 and R version 4.0.5. were used to conduct all analysis. The following R-Studio packages were required: dplyr, readxl, tidyr, ggplot2, openxlsx, forcats, scales, stringr, corrplot, patchwork, cowplot, ggrepel and gridExtra

### Reference IgG immunoassays

To further validate the multiplex assay, a comparison was carried out using matched samples from healthy French MI donors. Complementary antigen-specific serological tests were conducted to measure IgG levels within samples of the MI cohort, following the manufacturer’s guidelines. This validation was performed for a subset of ten antigens ^22^.

Anti-HBs and anti-HBc IgGs were measured on the Architect automate (CMIA assay, Abbott). Anti-CMV IgGs were measured by CMIA using the CMV IgG kit from Beckman Coulter on the Unicel Dxl 800 Access automate (Beckman Coulter). Anti-measles, anti-mumps, and anti-rubella IgGs were measured using the BioPlex 2200 MMRV IgG kit on the BioPlex 2200 analyzer (Bio-Rad). Anti-Toxoplasma *gondii* IgGs were measured using the BioPlex 2200 ToRC IgG kit on the BioPlex 2200 analyzer (Bio-Rad). Anti-influenza A IgGs were measured by ELISA using the NovaLisa IgG kit from NovaTec (Biomérieux) that explores responses to grade 2 H3N2 Texas 1/77 strain. In all cases, the manufacturers established the criteria for serostatus definition (positive, negative, or indeterminate).

### Statistical analysis

To determine sero-positivity status it is necessary to select a cut-off for each measured antibody response. Due to substantial variation in the epidemiology of the studied pathogens, it was not possible to use a single statistical method for selecting a sero-positivity cut-off. We instead developed a decision tree based on prior knowledge of pathogen exposure or vaccination, and whether the data followed a bimodal distribution (Figure 1).

**Figure 1:**
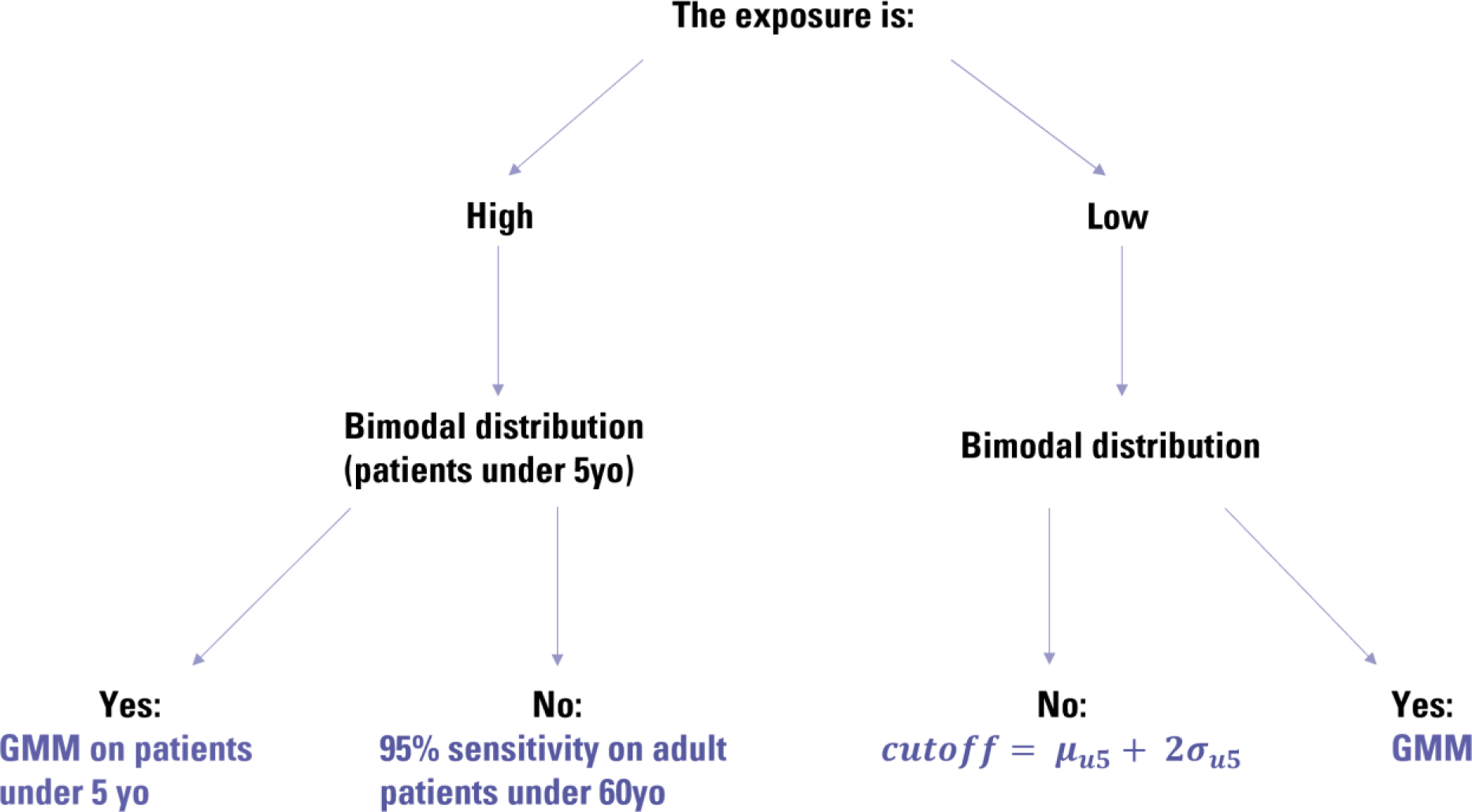
Decision tree used to choose between the different methods available to determine the cut-off discriminating between the seropositive and seronegative patients.

The decision of low or high exposure was based on prior epidemiological knowledge for that pathogen. In cases of low exposure, if a visual examination of the data indicated that log antibody levels followed a bimodal distribution, we applied a Gaussian Mixture Model (GMM). This method was applied only to the Cytomegalovirus antigen.

In cases of low exposure when the data did not follow a bimodal distribution, we analysed data from children <5 years. We assume that when exposure is low, the majority of young children are unexposed and seronegative. A cut-off was selected based on the mean plus two standard deviations. For normally distributed data, this corresponds to specificity = 97.7%. This method was applied to the Hepatitis A, HEV ORF2, HPV16 and HPV18 antigens.

In cases of high exposure, we examined log antibody levels in children <5 years. If the data followed a bimodal distribution, we applied a GMM. This method was applied to data from Adenovirus T3 and T5, HKU1 Spike, OC43 Spike, Echovirus, NL63 S, 229E Spike, Measles nucleoprotein, Measles lysate, Rubella, Norovirus GII.4 VP1, RSV B, Epstein-Barr virus, Tetatuns Toxin (NIBSC), and Varicell-Zoster antigens.

When there was no evidence for a bimodal distribution, we focused on data from individuals aged 18 – 60 years. We assume that when exposure is high, the majority of adults are exposed and seropositive. We exclude adults >60 years because of the possibility of immunosenescence. In this framework, it was not possible to control specificity, so we instead select a cut-off corresponding to sensitivity = 95%. As the data were not always Normally distributed, we do not attempt to calculate means and standard deviation, but instead calculate the cut-off based on the 5% percentile of the data. This method was applied to Bordella p. Toxin, OC43 NP, HKU1 nucleoprotein, NL63 nucleoprotein, Enterovirus CoxB3 VP1, Adenovirus T5, Mumps lysate, Norovirus GII. 6 VLP, RSV A, RSVglycoprotein G, Rhinovirus T1A lysate, Rotavirus VP7, Tetanus toxin (NA), Mumps, Rubella E2, 229E nucleoprotein, Mumps nucleoprotein, Influenza A, Bordetella p.FHA, ADE40, Norovirus GII.4 VLP antigens.

A Gaussian Mixture Model was used if the data were bimodally distributed. Formally, *Ab*_*i*_ denotes the antibody level (MFI) of patient *i* and *Ab*_*i*_ ∼ (1 − *θ*)*N*(*μ*_*neg*_, *σ*_*neg*_) + *θN*(*μ*_*pos*_, *σ*_*pos*_), with *θ* the prevalence of seropositive patients in this subpopulation, *μ*_*neg*_ and *μ*_*pos*_ the mean antibody levels of the seronegative and seropositive patients respectively and *σ*_*neg*_ and *σ*_*pos*_ their standard deviation. Thus, the model was defined by the following equation:

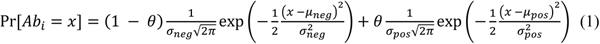

From which the estimated cut-off was derived as: *μ*_*neg*_ + 2*σ*_*neg*_.

The cut-off determined in MFI were then converted to RAU using the same method as for the samples.

To compare the seroprevalence rates between males and female, Wilcoxon statistical tests were performed using Rstudio.

## Results

### Vaccine preventable diseases panel

Antibody levels against vaccine preventable diseases from different age groups are shown in Figure 2. High antibody levels are detected in children <1 year old (newborns), consistent with the presence of maternal antibodies. When examining antigens such as Measles nucleoprotein or Rubella VLP, we not only observe the initial presence of antibodies but also their subsequent decline before the administration of the MMR vaccine, typically around 12 to 18 months of age.

**Figure 2:**
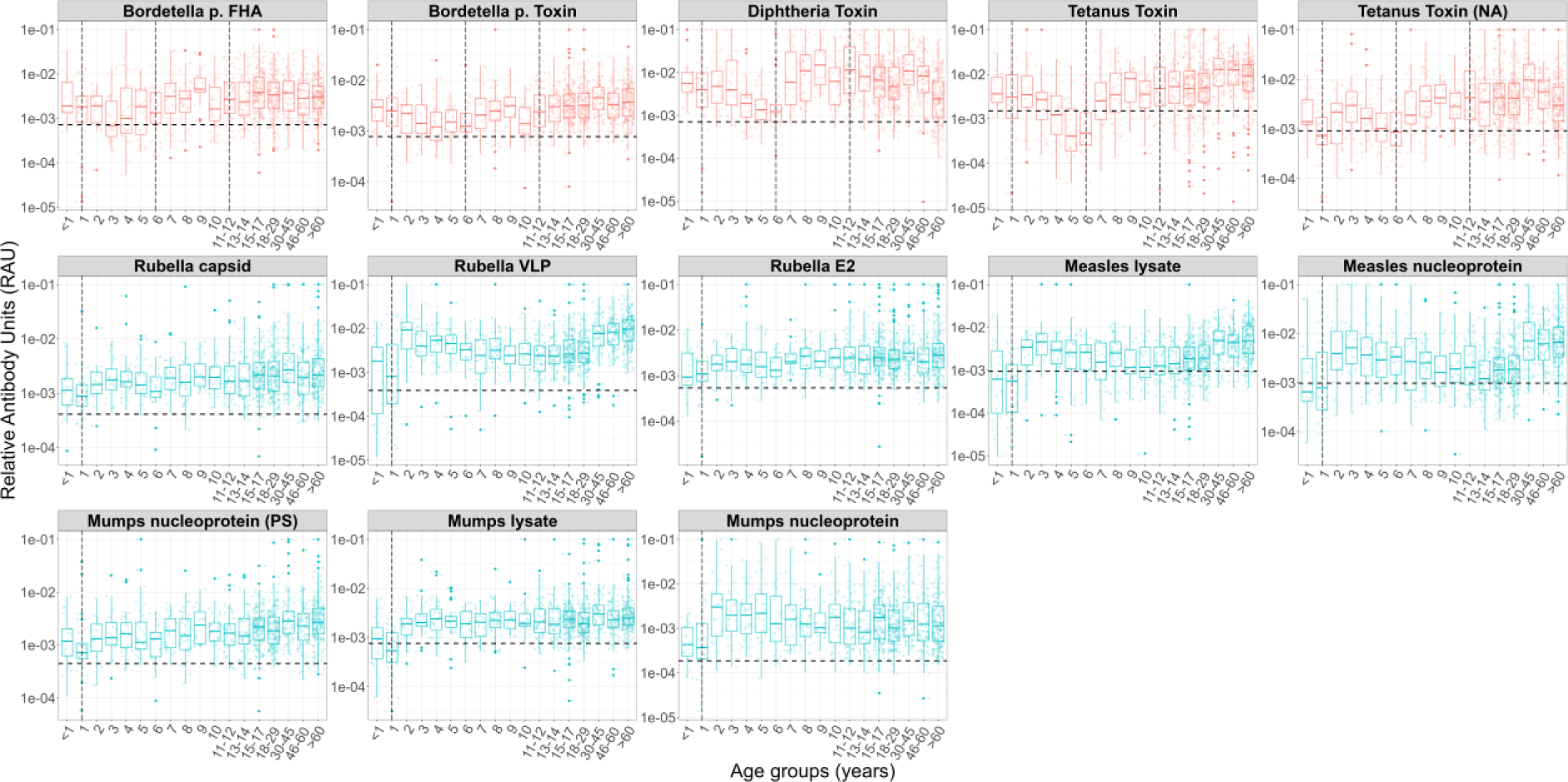
Variation in vaccine preventable disease antibody levels among different age groups. Relative Antibody Units (RAU) are calculated using the median fluorescence intensity (MFI) from a Luminex INTELLIFLEX. The horizontal dotted lines represent the antigen-specific cut-off for seropositivy. Vertical dotted lines denote specific recommended vaccination time points. Antigens were colour-coded, each representing a specific vaccine, red for Diphtheria, Tetanus, and Pertussis (DTaP) vaccine; blue for Measles, Mumps, Rubella (MMR) vaccine.

Although we do not have data on which individuals were vaccinated, a notable increase in antibody titers is observed following the recommended timing of vaccine doses. This is particularly evident for *Bordetella pertussis* toxin and FHA antigens, where multiple booster doses are administrated over several years. Following the booster dose there is a decline in antibody levels; however, antibody levels remain above the initial pre-vaccination baseline. A similar pattern of boosting followed by waning is observed for Tetanus and Diphtheria toxin, which are administered simultaneously as part of the DTaP vaccine. Furthermore, high antibody levels in older individuals are consistent with vaccination guidelines, which advise for a DTaP booster dose every 10 years or after experiencing an injury as a precautionary measure. This phenomenon is also observed for antigens such as Measles lysate, Measles nucleoprotein, and Rubella VLP, where a rise in antibody titers is observed during adulthood, despite no administration of booster doses.

The majority of individuals pass the established seropositivity cut-off for mandatory vaccines (e.g. MMR vaccine and DTaP vaccine) from a very young age. However, for the two HPV antigens included within the panel, we refrain from presenting further analyses of seroprevalence, due to a lack of confidence in the quality of our HPV serological assay. Age-stratified results have been reported in the Supplementary Fig 2.

### Common viral infections multiplex panel

As shown previously, newborns exhibit high levels of antibodies, indicating the transfer of antibodies acquired during the mother’s life. For diseases where vaccination is unavailable for infants and toddlers, the decline of maternal antibodies during the first year of life is apparent. This decrease is particularly evident for Adenovirus and VZV. (Figure 3)

**Figure 3:**
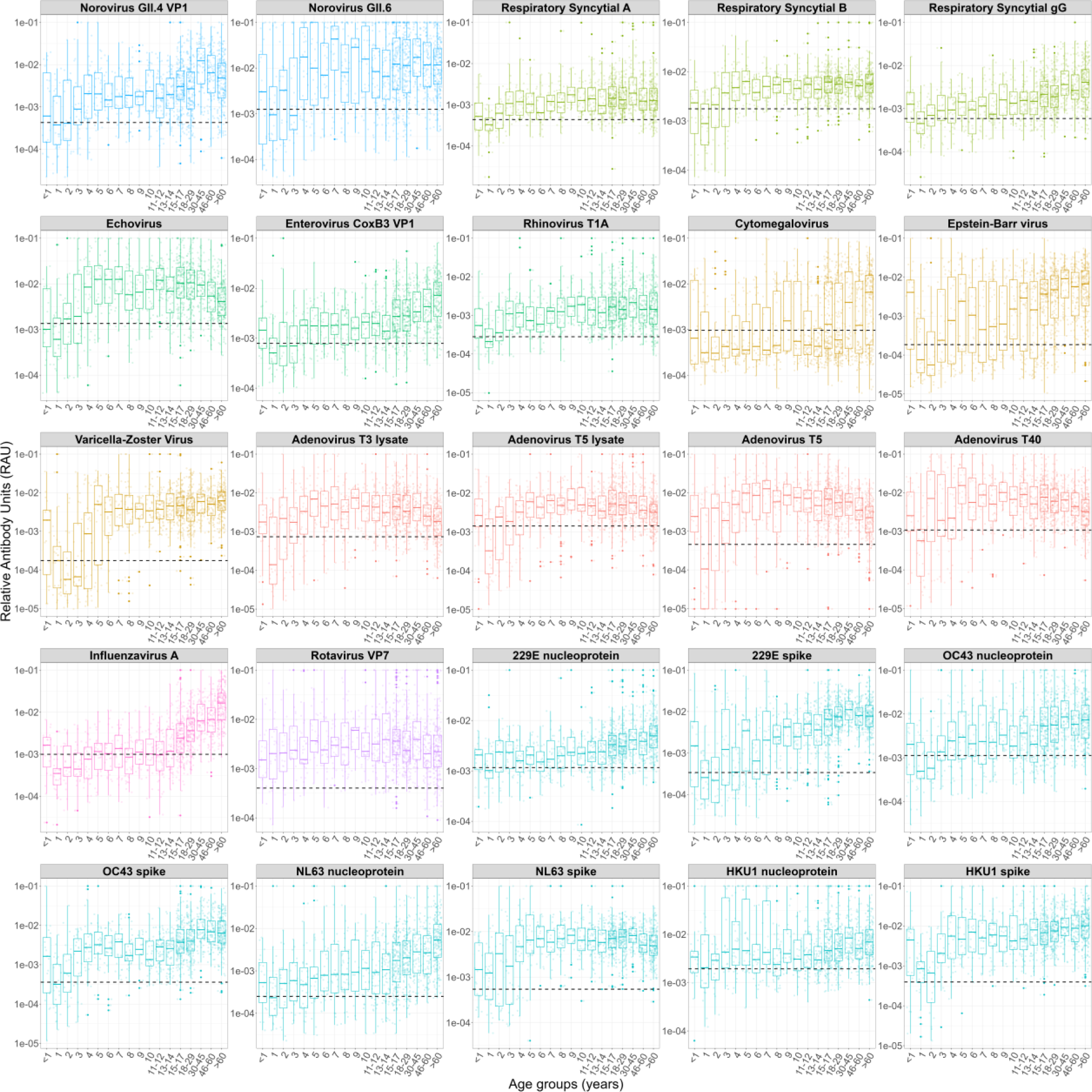
Age-related variation in antibody levels against common viral infections. RAUs have been generated using the MFIs measurements. The horizontal dotted lines represent the antigen-specific cut-offs for seropositivity. Antigens were colour-coded for clarity, with each distinct colour corresponding to a specific virus family or genus.

For gastro-intestinal viruses, there was a noticeable rise in antibody levels around two years of age, implying early exposure to these viruses. This pattern, characterized by the decline in temporary immunity conferred by the mother and the subsequent rise in antibodies following the first infection, is notably pronounced with the different antigens of Adenovirus. In contrast, a distinct pattern unfolds with the Varicella-Zoster virus, revealing a sharp increase in antibody levels around 4 years of age. This suggests that the first exposure to this virus occurs later in life, possibly in environments such as school or daycare.

For respiratory viruses, notably RSV, the majority of individuals exhibit antibody levels above the seropositivity threshold. Indicating a first infection typically occurring before 2-3 years of age and continuous exposure throughout life, maintaining consistently high levels of antibodies.

Although there is no discernible age-related trend or early exposure observed for Influenza A or CMV, an increase in antibody levels among adults is observed suggesting continued exposure in older age groups. For Influenza A, a significant rise in antibody titer among the elderly is revealed, likely influenced by annual influenza vaccination campaigns specifically tailored to target the older demographic.

### Multiplex assay validation

Our assay underwent validation using clinical-grade ELISA assays by evaluation of the correlation in measured antibody levels between the two assays. ELISA assays were conducted on all samples from the Milieu Intérieur cohort on a subset of ten antigens. This subset included four vaccine preventable diseases: Rubella, Mumps, Measles, and Hepatitis B as well as common persistent/recurrent viruses: CMV, EBV, VZV, Influenza A, along with the parasite *Toxoplasma gondii*. In contrast to our assay, some commercially available ELISA tests lack information regarding the specific antigen target, however we have access to raw quantitative data along with the clinical interpretation of the serological status (either seropositive, seronegative or undefined/grey zone).

Strong correlations were observed for antigens such as Rubella, Measles, and VZV with approximately 93.7%, 91.9% and 93.3% of individuals classified as seropositive (showed in green) with the reference assays (Figure 4). This aligned closely with our in-house assay, which showed a seroprevalence of 97.8%, 90.3%, and 99.8% for each virus, respectively. Despite a high degree of correlation of measured antibody levels between assays for both Rubella and VZV, the estimated seroprevalence using the multiplex assay was substantially higher than when using ELISA. We attribute this to the selection of a more sensitive sero-positivity cut-off for the multiplex assay. For the remaining antigens the correlation was less pronounced. For EBV, a large number of samples reached the upper limit of the assay, whereas with our multiplex assay, they exhibited a broader range of RAU values, which results in the appearance of a weaker correlation. Lastly, the results for Toxoplasmosis are inconclusive, as no correlation was observed. Due to a lack of confidence in the quality of our serological assay for Toxoplasmosis, we refrain from reporting results for this pathogen.

**Figure 4:**
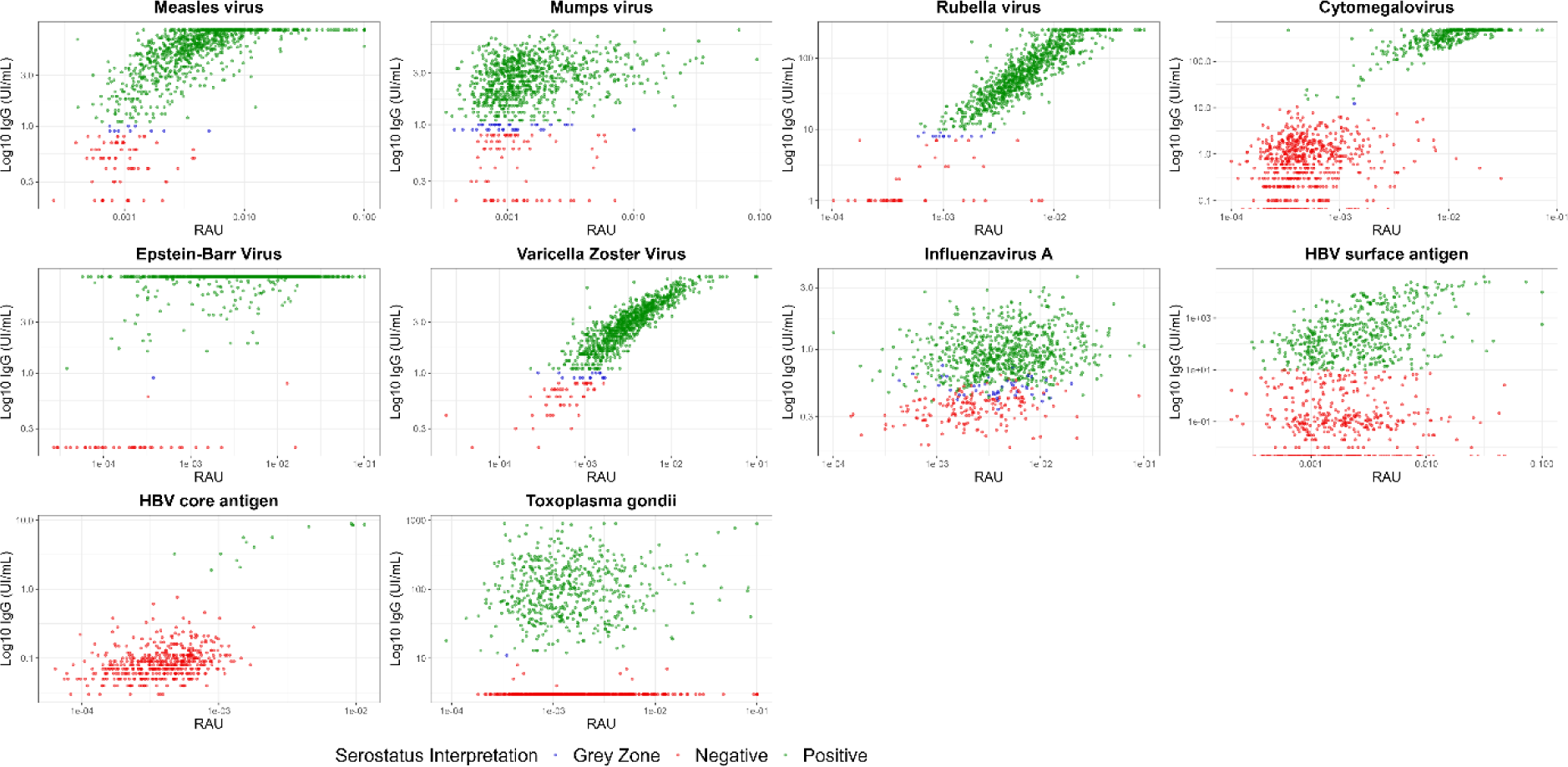
Correlation between serological data from two immunoassays. Comparison of the serological data generated by the in-house Luminex INTELLIFLEX assay and those from clinical-grade ELISA assays. The x-axis displays relative antibody units (RAU), while the y-axis represents the unit from the various reference tests. Serological interpretation of the clinical-grade assays is showed, distinguishing between negatives (red), positives (green) and indetermined (blue).

### Seroprevalence across diverse age groups in a population

After establishing antigen cut-offs using statistical models, the seroprevalence was calculated within the Seroped and Milieu Intérieur cohorts, both representing the healthy French population. The cohorts differed in their composition; the first included newborn to elderly individuals, and the second exclusively adults. To facilitate analysis, we categorized the population into four distinct groups: young children (under 5 years old), older children and teenagers (5 – 18 years old), adults from the Seroped study, and adults from Milieu Intérieur.

Seroprevalence for both mandatory and recommended vaccines in France is presented (Figure 5). Seroprevalence rates for mandatory vaccines are consistently high across all age groups and antigens. Among adults, the seroprevalence varies from 88.4% to 100% depending on antigens, indicating high vaccination coverage. Importantly, there is no evidence of seroreversion for any pathogen, implying that the administered vaccines effectively induce an immune response and maintains stable antibody levels throughout an individual’s life.

**Figure 5:**
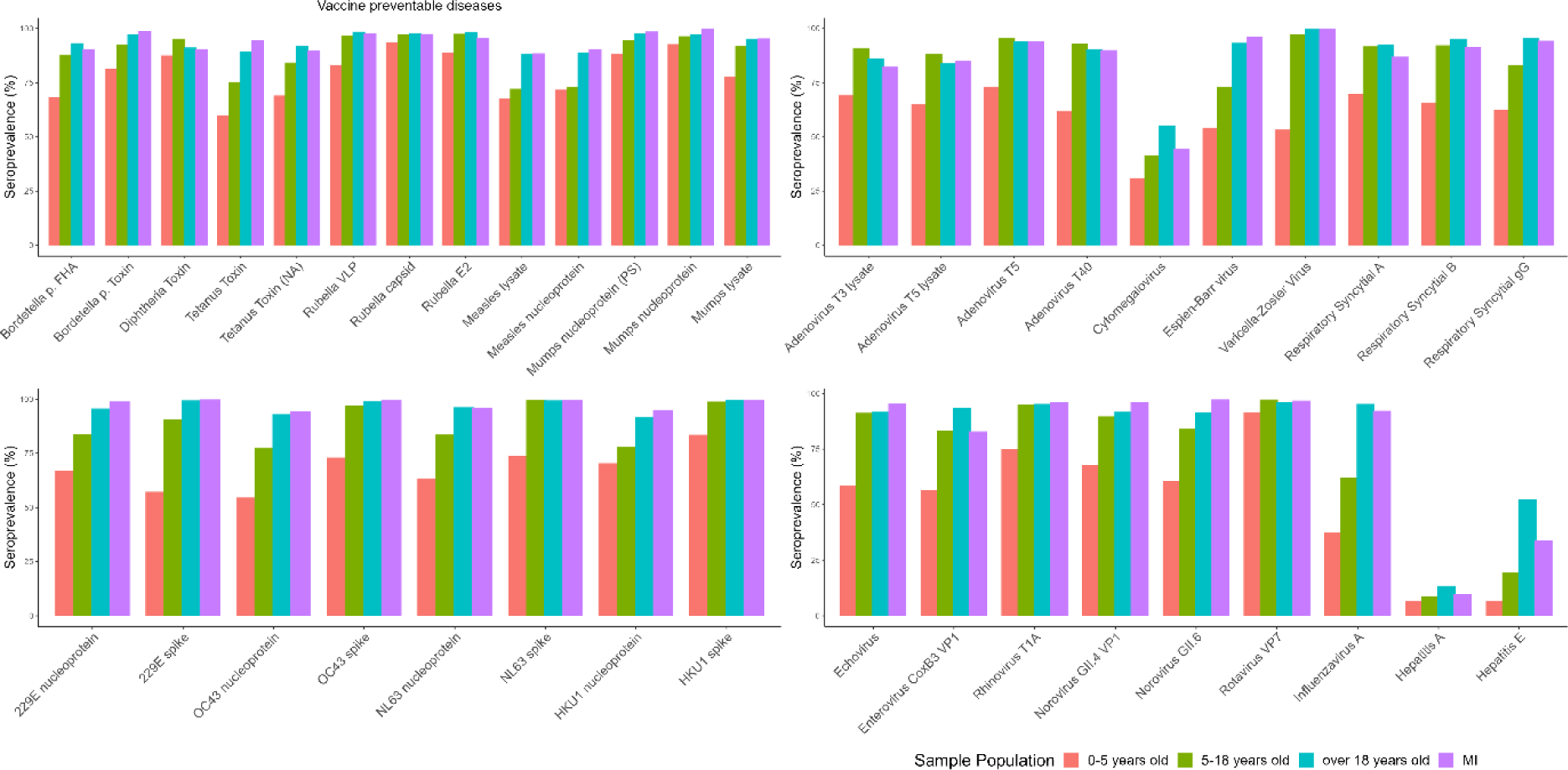
Seroprevalence across various age groups for vaccine-preventable diseases and common viral infections. Both cohorts are included, with the Seroped cohort divided into infants and toddlers (in red), children and adolescents (in green), adults (in blue), and all adults from the Milieu Intérieur study (in purple).

For pathogens in this study that target the respiratory or intestinal systems, seroprevalence remains high in all adults, with similar trends observed between vaccine-preventable diseases and high-prevalence viruses. Children under 5 years old exhibit varying seroprevalence rates, ranging from 30.9% for CMV to 98.7% for Rotavirus, indicating diverse virus exposure levels during early childhood. In contrast, adolescents and adults display seroprevalence ranging from 41.2% to 55.2% for CMV, reaching 100% for HKU1, reflecting ongoing exposure and long-term immunity.

The panel includes a dedicated section on hepatitis viruses; however, the seroprevalence rates for these antigens remain relatively low compared to other viral infections. The observed seroprevalence among adults in the MI cohort was 9.9% for Hepatitis A and 33.9% for Hepatitis E. In adults in the SeroPed study, the observed seroprevalence was 13.5% for Hepatitis A and 52.4% for Hepatitis E. We did not calculate the seroprevalence of Hepatitis B and Hepatitis C because we did not expect to observe enough positive antibody responses in our samples to allow accurate calculation of a cut-off. Supplementary Fig. 1 presents age-stratified results for all Hepatitis antigens comprised in our panel.

### Gender-specific seroprevalence

An analysis of seroprevalence based on gender was undertaken using data from the Seroped cohort (Figure 6). For most pathogens, no significant difference in seroprevalence between genders was observed (p-value > 0.05). However, a significantly higher seroprevalence among men for Tetanus has been shown. Seroprevalence rates for Tetanus Toxin were 75% among women and 82.7% among men (p-value = 0.002889), and 81.5% among women and 88.9% among men for Tetanus Toxin (NA) (p-value = 0.001096). The observed difference in seroprevalence between the two genders may be attributed to a higher incidence of accidents among men. Thereby an increased probability of receiving a tetanus booster, leading to higher antibody levels^23^. However, when we analyzed the quantitative antibody level to tetanus, we did not observe a significant difference. The absence of gender-based variation among the other pathogens may be attributed to similar exposure patterns and comparable susceptibility to infections, along with a harmonized immune response across both genders.

**Figure 6:**
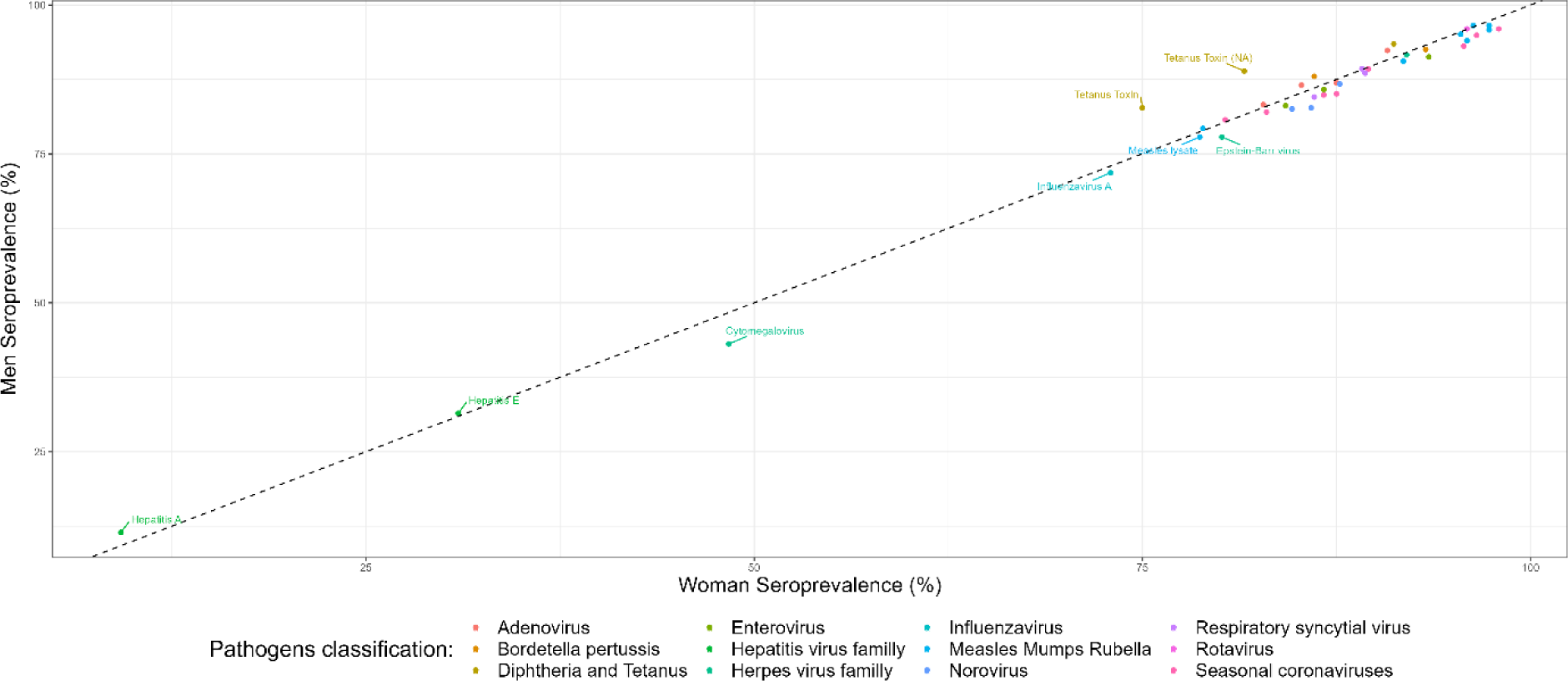
Correlation of seroprevalence between men and woman in the Seroped cohort. Antigens were color-coded for clarity, with each distinct color corresponding to a specific virus family, genus or group.

## Discussion

In this study, we demonstrate how a laboratory-based multiplex pathogen assay can discern variations in antibody titers across age groups within healthy populations, establish sero-positivity cut-offs, and ascertain seroprevalence. Additionally, it enables us to better understand the effects of vaccines and booster doses, provides insights into the transfer of passive immunity to infants through maternal antibodies, and assesses the gradual waning of antibodies over time.

The seroprevalence profiles of vaccine-preventable diseases (Figure 5), aligns closely with the high vaccination coverage rates for mandatory vaccines in France, reaching approximately 95%. This is consistent with estimates from the World Health Organization and UNICEF regarding national immunization coverage, indicating a 96% coverage for the complete DTaP vaccine schedule and around 90% coverage for MMR vaccines in France. The correlation between seroprevalence and vaccination coverage underscores the effectiveness of the immunization programs.^24^

The observed high antibody levels in newborns indicate the presence of maternal antibodies transferred from the mother, which play a vital role in providing temporary immunity to the newborn^25^. The lifespan of maternal antibodies in the infant’s system varies, typically lasting approximately 6 to 9 months^26,27^. This passive immunity wanes over time, and infants gradually develop their own immune responses. Antibody titers then increase with age, after repeat infections, especially evident with RSV and Adenovirus, suggesting early initial exposure typically around 2-3 years of age and re-infection occurring throughout life^28^. The timing disparity between certain infections may be associated with their distinct transmission dynamics or seasonal patterns^29^. RSV, transmitted via respiratory droplets, tends to infect infants during its seasonal peaks, whereas varicella, transmitted mainly through direct contact (and respiratory droplets), may manifest later in childhood as social interactions increase with age^30^. As observed with our assay, an increase in varicella antibody levels occurs around the age of 4 (Figure 3), confirming reports of first exposure typically taking place in environments such as school or daycare^31^. On the other hand, high rates of seroconversion for CMV were observed during adulthood, raising questions about the factors that could prevent some individuals from becoming seropositive upon later-life exposure, despite the high prevalence of the virus within the population^32^. Nevertheless, our seroprevalence estimates align with previously reported estimates from the UK, indicating a CMV seroprevalence rate of approximately 50% among European adults^5^.

Across all age groups, there is a consistent high seroprevalence for endemic viruses, suggesting widespread exposure from a young age and frequent reinfection throughout life, contributing to a balance between increased immunity from new infections (seroconversion) and declining antibody levels (seroreversion)^33^. Immunosenescence, the age-related decline in the functionality of both innate and adaptive immune responses, results in a reduction of antibodies levels in older individuals, as observed with certain diseases such as diphtheria (Figure 2) where antibody production diminishes with age.

Our findings for Hepatitis A and E are consistent with those of other research teams, showing low seroprevalence rates for these viruses among adults, reaching, 9.9%, and 33.9%, respectively ^34–37^. These results align with existing studies, such as the assessment of Hepatitis A seroprevalence in French children and adolescents, where the overall seroprevalence was 5%^38^. When analyzing our data for individuals under 18 years old in the Seroped cohort, we observe a slightly elevated seroprevalence of 6.6% for Hepatitis A. Futhermore, our assay revealed a seroprevance of 13.5% among adults aged 18 year and over in the Seroped study, mirroring the results of a separate French study that reported a seroprevalence of 16.5% for adults^34^. In a meta-analysis of Hepatitis E seroprevalence in Europe, France appears to have a higher rate compared to its neighbors, with a prevalence of 32% in the general population^39^. This difference could be attributed to the foodborne transmission of the virus, especially through undercooked or raw meat. However, estimates of Hepatitis E seroprevalence vary widely in the literature.

We explored various approaches to validate our test. Ideally, well-characterized samples, including both negative and positive, would be used for validation; however this was not possible given the large number of pathogens studied. Alternatively, a second validation strategy involves the study of large cross-sectional cohorts enriched with epidemiological data. The Seroped study, where the serological data was age- and sex-stratified, enabled us to observe the variation in antibody levels across different age groups within the population. Subsequently, we validated our assay by assessing the consistency between pre- and post-vaccination antibody titers according to the established French vaccine schedule. Furthermore, we performed comparative analyses against reference or gold-standard tests. In this context, we employed the Milieu Intérieur cohort’s serological data (IgG levels and serological status interpretation) allowing a correlation analysis for 9 of the 47 antigens present in our panel. Beyond establishing a strong correlation between the two methods, this comparison reveals the higher sensitivity exhibited by our assay.

The developed assay has a number of limitations. Primarily, the absence of well-characterized samples or positive pool for each pathogen studied, has complicated certain steps, such as selecting the optimum antigen concentration to use during bead coupling. To overcome this obstacle, different concentrations of antigen-coupled beads were tested against a wide sample set, revealing the dynamic range of the assay. The use of a universal reference pool, prepared with serum from individuals exhibiting a high MFI response against a broad number of antigens, is a pragmatic and effective solution when you do not have a positive pool against all pathogens comprised in the panel. This reference pool not only effectively addressed inter-plate variations but also standardized results by converting MFI into RAU. Subsequently, the difficulty in obtaining verified negative control samples for globally endemic viruses that commonly infect young children was encountered. We addressed this challenge by fitting gaussian mixture models to samples from young children to identify positive and negative subpopulations. These limitations, although present during the initial development process, highlight areas for improvement to consider for the future.

The quality of the antigens had an important influence on overall assay performance. Low protein quality or stability can induce the degradation of the antigen coupled onto the bead or lead to inadequate refolding of the antigen. Additionally, distinct complications were observed depending on the nature of the antigens utilized, whether they were recombinant proteins, virus-like particles (VLPs), or lysates. When employing lysates, aggregation of beads occasionally appeared, leading to a reduction in the bead count during plate reading. VLPs, characterized by their larger-sized particles mimicking the virus structure, appeared to be more immunogenic, as indicated by the high MFI response. However, this heightened MFI could potentially be attributed to increased cross-reactivity with other viruses. Evaluating potential cross-reactivity between different antigens posed a challenge, as we would need large volume of specific anti-serum, which was not feasible during this study. Further tests should be preformed in order to understand and overcome possible cross-reactivity.

To conduct a more detailed evaluation of antibody kinetics, infections, and reinfection rates, a longitudinal study design would be more appropriate to better understand acquired immunity and vaccine effectiveness at the individual level. The identification of boosts in antibody levels between consecutive samples becomes feasible, thereby enabling the subsequent identification of infections and reinfections.

In the near future, we aim to overcome the previously reported limitations and expand the potential applications of our assay. It is contemplated to adapt this protocol to measure additional isotypes, as well as to conduct our assay using nasopharyngeal or saliva samples, as it has been demonstrated that specific mucosal IgA antibodies have a crucial role in early virus neutralization and long-term protection^40^. Further development of these protocols would provide us with a larger dataset enabling us to gain insights into the kinetics of antibodies and the role of different immunoglobulin isotypes post-immunization.

## Conclusion

This study addresses the challenges of serosurveillance of vaccine-preventable diseases and common viral infections through the development of a high-throughput multiplex serology assay, providing insights into natural and vaccine-acquired immunity. The findings not only contribute to a comprehensive understanding of immunization through vaccine dose effects but also successfully evaluate vaccine coverage, identifying potential gaps. Among infectious diseases for which no vaccine is available, this tool enables us to study the role of maternal antibodies in providing temporary immunity. Additionally, it allows us to estimate the time of the first infection and evaluate the decline of antibodies over time. Therefore, the development of serosurveillance tools are essential to monitor the immune status of the population and design effective vaccination strategies to prevent outbreaks and improve global health.

## Data Availability

All data produced in the present study are available upon reasonable request to the authors

## Author contributions

EB developed the assay, analysed samples, and wrote the first draft of the manuscript. GB analysed the data. LG, FD and SP supported assay development. AF, DD and LQM provided access to samples. MW designed the study.

## Competing interests

All authors report that they have no conflicts of interest.

## Acknowledegments

This work was supported by the the French government’s “Integrative Biology of Emerging Infectious Diseases” (Investissement d’Avenir grant ANR-10-LABX-62-IBEID) and INCEPTION programs (Investissement d’Avenir grant ANR-16-CONV-0005); and the “URGENCE COVID-19” fundraising campaign of Institut Pasteur. Milieu Intérieur was funded by the French government’s Invest in the Future programme; reference ANR-10-LABX-69-01.

## Notes

### Competing Interest Statement

The authors have declared no competing interest.

### Funding Statement

This work was supported by the the French government: Integrative Biology of Emerging Infectious Diseases (Investissement d Avenir grant ANR-10-LABX-62-IBEID) and INCEPTION programs (Investissement d Avenir grant ANR-16-CONV-0005), and the "URGENCE COVID-19" fundraising campaign of Institut Pasteur. Milieu Interieur was funded by the French governments Invest in the Future programme: reference ANR-10-LABX-69-01.

### Author Declarations

Milieu Interieur The Comite de Protection des Personnes: Ouest 6 of the French Agence Nationale de Securite du Medicament gave ethical approval for this work. The clinical study was approved by the Comite de Protection des Personnes Ouest 6 on June 13, 2012, and by the French Agence Nationale de Securite du Medicament on June 22nd, 2012. The study is sponsored by Institut Pasteur (Pasteur ID RCB Number: 2012 A00238 35) and was conducted as a single center study without any investigational product. The original protocol was registered under ClinicalTrials.gov (study# NCT01699893). The samples and data used in this study were formally established as the Milieu Interieur biocollection (NCT03905993), with approvals by the Comite de Protection des Personnes Sud Mediterranee and the Commission Nationale de l'Informatique et des Libertes (CNIL) on April 11, 2018. SeroPed The Clinical Research Coordination Office of Institut Pasteur waived ethical approval for this work. The samples analysed in this study were leftovers from routine medical blood sample processing in French hospital laboratories. They were processed in accordance with existing regulations and guidelines of the French Commission for Data Protection (Commission Nationale de l'Informatique et des Liberties). Sera were completely anonymous, and it was not possible to return to individual patients files. According to the French law, no informed consent is required for processing such samples.

## References

1. A prospective analysis of the Ab response to Plasmodium falciparum before and after a malaria season by protein microarray | PNAS. https://www.pnas.org/doi/10.1073/pnas.1001323107.

2. Shrock, E. L., Shrock, C. L. & Elledge, S. J. VirScan: High-throughput Profiling of Antiviral Antibody Epitopes. Bio-Protoc. 12, e4464 (2022).

3. Van Gageldonk, P. G. M., Van Schaijk, F. G., Van Der Klis, F. R. & Berbers, G. A. M. Development and validation of a multiplex immunoassay for the simultaneous determination of serum antibodies to Bordetella pertussis, diphtheria and tetanus. J. Immunol. Methods 335, 79–89 (2008).

4. Rosado, J. et al. Multiplex assays for the identification of serological signatures of SARS-CoV-2 infection: an antibody-based diagnostic and machine learning study. Lancet Microbe 2, e60–e69 (2021).

5. Mentzer, A. J. et al. Identification of host-pathogen-disease relationships using a scalable multiplex serology platform in UK Biobank. Nat. Commun. 13, 1818 (2022).

6. Sheikh, S. et al. A report on the status of vaccination in Europe. Vaccine 36, 4979–4992 (2018).

7. den Hartog, G., van Binnendijk, R., Buisman, A.-M., Berbers, G. A. M. & van der Klis, F. R. M. Immune surveillance for vaccine-preventable diseases. Expert Rev. Vaccines 19, 327–339 (2020).

8. Alter, S. J., Bennett, J. S., Koranyi, K., Kreppel, A. & Simon, R. Common Childhood Viral Infections. Curr. Probl. Pediatr. Adolesc. Health Care 45, 21–53 (2015).

9. Teoh, Z. et al. Burden of Respiratory Viruses in Children Less Than 2 Years Old in a Community-based Longitudinal US Birth Cohort. Clin. Infect. Dis. 77, 901–909 (2023).

10. Jackson, D. J. The role of rhinovirus infections in the development of early childhood asthma. Curr. Opin. Allergy Clin. Immunol. 10, 133–138 (2010).

11. Guarino, C. et al. Development of a quantitative COVID-19 multiplex assay and its use for serological surveillance in a low SARS-CoV-2 incidence community. PloS One 17, e0262868 (2022).

12. Al-Tawfiq, J. A. et al. Surveillance for emerging respiratory viruses. Lancet Infect. Dis. 14, 992–1000 (2014).

13. Hao, R. et al. Surveillance of emerging infectious diseases for biosecurity. Sci. China Life Sci. 65, 1504–1516 (2022).

14. Iwane, M. K. et al. Population-Based Surveillance for Hospitalizations Associated With Respiratory Syncytial Virus, Influenza Virus, and Parainfluenza Viruses Among Young Children. Pediatrics 113, 1758–1764 (2004).

15. Galanti, M. et al. Rates of asymptomatic respiratory virus infection across age groups. Epidemiol. Infect. 147, e176 (2019).

16. Gardner, G., Frank, A. L. & Taber, L. H. Effects of social and family factors on viral respiratory infection and illness in the first year of life. J. Epidemiol. Community Health 38, 42–48 (1984).

17. Ferson, M. J. Infections in day care. Curr. Opin. Pediatr. 5, 35–40 (1993).

18. Churchill, R. B. & Pickering, L. K. Infection control challenges in child-care centers. Infect. Dis. Clin. North Am. 11, 347–365 (1997).

19. Slifka, M. K., Antia, R., Whitmire, J. K. & Ahmed, R. Humoral immunity due to long-lived plasma cells. Immunity 8, 363–372 (1998).

20. Woudenberg, T., et al. Humoral Immunity to SARS-CoV-2 and Inferred Protection from Infection in a French Longitudinal Community Cohort. (2022).

21. Thomas, S., et al. *The Milieu Intérieur* study — An integrative approach for study of human immunological variance. Clin. Immunol. 157, 277–293 (2015).

22. Olin, A. et al. A systematic investigation into the non-genetic and genetic factors affecting the human anti-viral antibody repertoire. 2023.11.07.23298153 Preprint at 10.1101/2023.11.07.23298153 (2023).

23. Gergen, P. J. et al. A Population-Based Serologic Survey of Immunity to Tetanus in the United States. N. Engl. J. Med. 332, 761–767 (1995).

24. Bechini, A. et al. Childhood vaccination coverage in Europe: impact of different public health policies. Expert Rev. Vaccines 18, 693–701 (2019).

25. Dolatshahi, S. et al. Selective transfer of maternal antibodies in preterm and fullterm children. Sci. Rep. 12, 14937 (2022).

26. Madani, G. & Heiner, D. C. Antibody transmission from mother to fetus. Curr. Opin. Immunol. 1, 1157–1164 (1989).

27. Grindstaff, J. L., Brodie, E. D. & Ketterson, E. D. Immune function across generations: integrating mechanism and evolutionary process in maternal antibody transmission. Proc. R. Soc. B Biol. Sci. 270, 2309–2319 (2003).

28. Lu, G. et al. Large-scale seroprevalence analysis of human metapneumovirus and human respiratory syncytial virus infections in Beijing, China. Virol. J. 8, 62 (2011).

29. Moriyama, M., Hugentobler, W. J. & Iwasaki, A. Seasonality of Respiratory Viral Infections. Annu. Rev. Virol. 7, 83–101 (2020).

30. Azzari, C. et al. Epidemiology and prevention of respiratory syncytial virus infections in children in Italy. Ital. J. Pediatr. 47, 198 (2021).

31. Viner, K. et al. Transmission of varicella zoster virus from individuals with herpes zoster or varicella in school and day care settings. J. Infect. Dis. 205, 1336–1341 (2012).

32. Scepanovic, P. et al. Human genetic variants and age are the strongest predictors of humoral immune responses to common pathogens and vaccines. Genome Med. 10, 59 (2018).

33. De Thoisy, A. et al. Seroepidemiology of the Seasonal Human Coronaviruses NL63, 229E, OC43 and HKU1 in France. Open Forum Infect. Dis. 10, ofad340 (2023).

34. Lagarde, E., Joussemet, M., Lataillade, J. J. & Fabre, G. Risk factors for hepatitis A infection in France: drinking tap water may be of importance. Eur. J. Epidemiol. 11, 145–148 (1995).

35. Mansuy, J.-M. et al. Hepatitis E Virus Antibodies in Blood Donors, France. Emerg. Infect. Dis. 17, 2309–2312 (2011).

36. Kwon, S. Y. & Lee, C. H. Epidemiology and prevention of hepatitis B virus infection. Korean J. Hepatol. 17, 87–95 (2011).

37. Hudu, S. A. et al. Isolated hepatitis B core antibody positive among vaccinated cohort in Malaysia. Ann. Saudi Med. 33, 591–594 (2013).

38. Faillon, S. et al. Impact of travel on the seroprevalence of hepatitis A in children. J. Clin. Virol. 56, 46–51 (2013).

39. Hartl, J. et al. Hepatitis E Seroprevalence in Europe: A Meta-Analysis. Viruses 8, 211 (2016).

40. Sterlin, D. et al. IgA dominates the early neutralizing antibody response to SARS-CoV-2. Sci. Transl. Med. 13, eabd2223 (2021).

